# The Gender Gap in Leading Medical Journals - a Computational Audit

**DOI:** 10.1101/2022.12.21.22283801

**Authors:** Oscar Brück

## Abstract

**Background:** Publication track record can impact careers of researchers. Therefore, monitoring gender representation in medical research is required to achieve equity in academia.

**Methods:** We gathered bibliometric data on original research articles published between 2010 and 2019 in *The New England Journal of Medicine, Nature Medicine, Journal of the American Medical Association, The BMJ*, and *The Lancet* using the Web of Science indexing database. We associated publication and citation frequency with author gender, count, and institute affiliation, and research keywords.

**Findings:** We analyzed 10,558 articles and found that women published and were cited less than men. There were fewer women as senior (24.8%) than leading authors (34.5%, p<0.001). The proportion of female authors varied by country with 9.1% last authors from Austria, 0.9% from Japan, and 0.0% from South Korea. The gender gap decreased longitudinally and faster for last (−24.0 articles/year, p<0.001) than first authors (−14.5 articles/year, p=0.024). The trend varied by country and even increased in China and Israel. Author count was associated with higher citation count (*R* 0.46, p<0.001) as well as with male first (n=11 vs. n=10, p<0.001) and last authors (n=11 vs. n=10, p<0.001). We also discovered that usage of research keywords varied by gender, and it partly accounted for the difference in citation counts by gender.

**Interpretation:** Gender representation has increased both at the leading and senior author levels although with country-specific variability. The study frame can be easily applied to any journal and time period to monitor changes in gender representation in science.

## BACKGROUND

Gender equity in medical research refers to the equal representation, recognition, and valuation of all researchers regardless of their gender. Women and other underrepresented groups continue to face barriers in diverse research fields. The proportion of graduating female medical students is equal to that of men^1^. Yet, publications are more frequently authored^2,3^, peer-reviewed^4^, and editorially evaluated by men^5,6^. Based on a report by the Association of American Medical Colleges, the proportion of women faculty had increased to 41% in 2019, while their proportion as department chairs remained at 18%^7^.

Abrogating the gender gap in all levels of academia are official priorities of the National Institutes of Health^8^ and the European Commission^9^. Promoting gender equity can lead to more representative and balanced research. In addition, it can help to reduce bias and discrimination in academic careers by ensuring that all researchers are treated fairly and given equal opportunities to succeed.

We recently discovered previously undocumented Anglocentric bias in leading medical journals based on publication counts and their citation frequency^10^. We reasoned that gender underrepresentation could be studied with a similar approach and explore publication differences in research fields, scientific journals, and collaboration scope.

## METHODS

### Data collection

For this study, we collected data from five medical journals that were ranked highest in the Journal Citation Reports 2022 and published primarily original articles (Fig. 1A). These were *The New England Journal of Medicine* (NEJM), *Nature Medicine* (NatMed), *Journal of the American Medical Association* (JAMA), *The BMJ*, and *The Lancet*. We included all original articles published from 2010 to 2019 totaling to 10,558 articles. We excluded more recent publications to avoid bias related to the COVID-19 pandemic and to ensure equal opportunities for accumulating citations. We queried for articles in the Web of Science database by Clarivate Plc with the terms “(((SO=(NATURE MEDICINE OR LANCET OR NEW ENGLAND JOURNAL OF MEDICINE OR JAMA JOURNAL OF THE AMERICAN MEDICAL ASSOCIATION OR BMJ BRITISH MEDICAL JOURNAL)) AND DT=(Article)) AND PY=(2010-2019))”. This allowed us to download metadata for each article, including the names of the authors, the address of the corresponding author, and the total number of citations.

**Figure 1.**
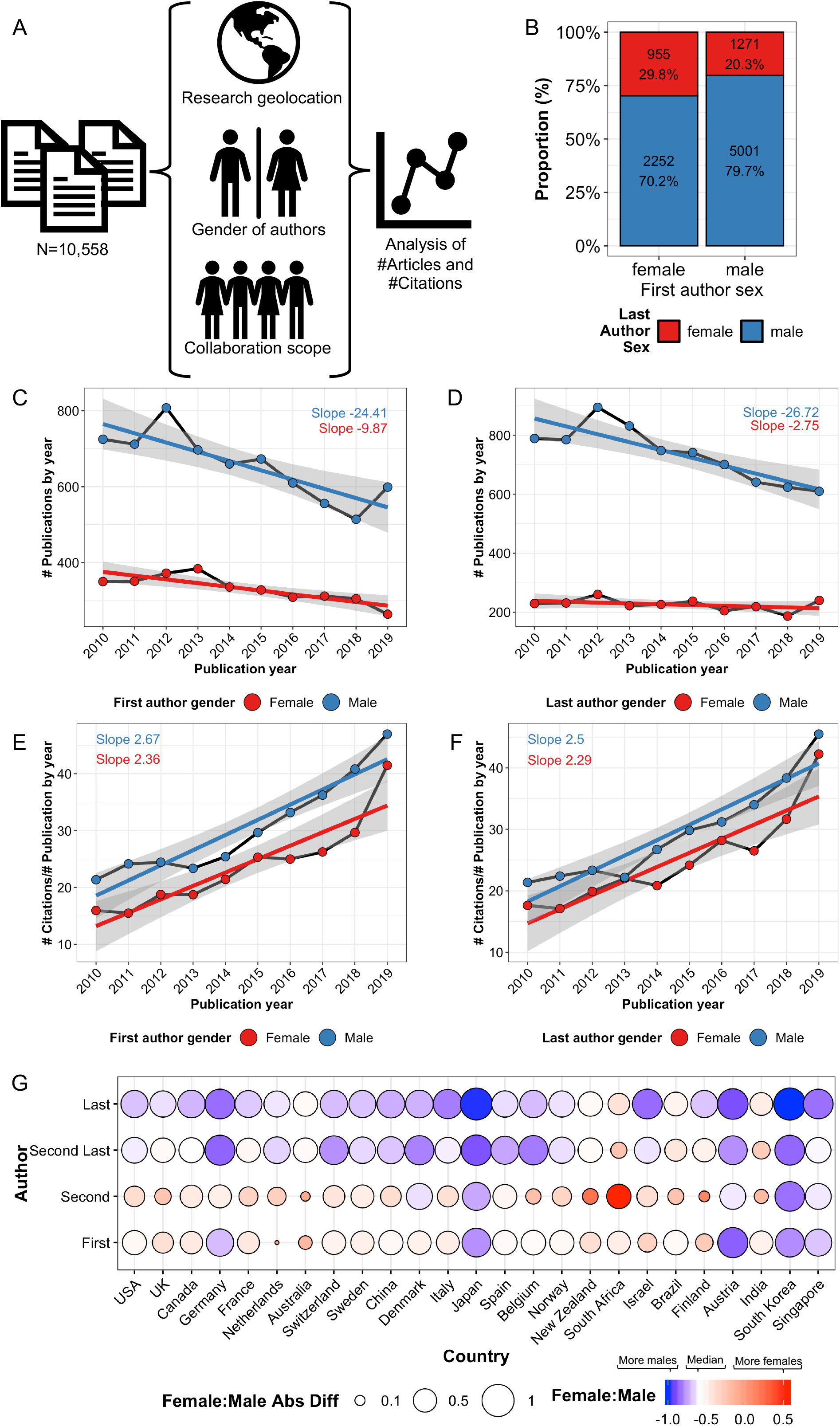
Gender gap in medical publishing. (A) Study design. (B) Bar plot on the association between first and last author gender. (C-D) Line plot and fitted linear regression for the number of publications and (E-F) number of yearly-averaged citations per publication by their publishing year and author gender. (G) Balloon plot on the association of the first, second, second last and last author gender distribution by the affiliation nationality of the corresponding author. The balloon color reflects the gender bias. To emphasize differences white color defines the median, i.e. men than women authors (red = more women, orange = balanced, blue = more men). The balloon size reflects the absolute deviation from a balanced gender distribution.

### Preprocessing variables of interest

We measured the impact of an article by its average citation count per year. To determine the number of authors, we summed the frequency of the semicolon “;” delimiter between author names and then added 1. For author names, only the initials of the forename were available for 538 first (5.1%), 529 second (5.0%), 780 second last (7.4%) and 572 last authors (5.4%). In total, we identified 7,558 unique forenames and defined the gender for 7,113 (94.1%) forenames with the genderizeR library based on the genderize.io database.

To determine the geographic location (latitude and longitude coordinates) of the primary institutes where the research was conducted, we applied the ggmap library and Google’s Geocoding API to the corresponding author’s address. If we were unable to successfully match the address, we geolocated the text-mined city and country of the address. The combined approach resulted in successful geolocation of 10,730 out of 10,732 total unique address (100.0%).

In the keyword analysis, we identified 21,820 unique keywords. As most of these were rarely used, we included only keywords employed in at least 20 articles (n=583, 2.7%).

### Statistical analysis

We used the Wilcoxon rank-sum test (unpaired, two-tailed) to compare two continuous variables and the Kruskal-Wallis test to compare three or more continuous variables. For categorical variables, we used the χ2 test. We adjusted p values using the Benjamini-Hochberg correction. To compare two linear regression slopes, we tested the significance of the interaction term using T-test. All statistical analyses and visualizations were conducted using R 3.5.1. using base, tidyverse, fastDummies, maps, reshape2, ggmap, data.table, countrycode, ggpubr, ggrepel, rstatix, ggdendro and dendextend libraries.

## RESULTS

### The gender gap in productivity is declining but not the gap in citation count

First, we interrogated how authoring patterns would differ between female and male researchers in original articles published in 2010-2019 in five leading medical journals (Fig. 1A). Publications with a male first author were two times and with a male last author three times as common compared to publications authored by a female first and last author, respectively (Table 1). In addition, publications with female first authors were 46.8% more likely to have a female last author compared to publications with a male first author (χ^2^ p<0.001, Fig. 1B).

**Table 1.** Association of gender authorship with publication metrics. Median and 25-75% interquartile ranges are reported.

| Authorship | Variable | Female | Male | P-value |
| --- | --- | --- | --- | --- |
| First | # Publications | 3311 | 6554 |  |
| First | # Citations | 22.0 [10.9-46.4] | 29.0 [13.6-60.9] | *** |
| First | # Authors | 10.0 [6.0-17.0] | 11.0 [6.0-18.0] | *** |
| Last | # Publications | 2256 | 7366 |  |
| Last | # Citations | 23.5 [11.0-52.4] | 28.2 [13.4-58.3] | *** |
| Last | # Authors | 10.0 [5.0-17.0] | 11.0 [6.0-18.0] | *** |

Temporally, the publication count has gradually converged between women and men (Fig. 1C). We fitted a linear regression for publication count using publication year and gender as covariate. The interaction term of the regression model was significant (coefficient -14.5, p=0.024) indicating that the yearly decline of first authorships for male researcher was higher in comparison to female researchers. The decline was even more distinct for last authors (coefficient -24.0, p<0.001 for the interaction term; Fig. 1D). Collectively, the gender gap in top medical research has declined with an average 14.5 publications/year for first authors and 24 publications/year for senior authors.

Female-authored articles gathered also fewer citations (Table 1). The median citation frequency by publication increased in line with rising medical journal impact factors (Fig. 1E-F). However, the ascent was equal between male and female researchers for both first (p=0.53 of the interaction term between publication year and gender; Fig. 1E) and last authors (p=0.67; Fig. 1F) implying no convergence in accumulated citations.

### The gender gap in productivity varies by country

The proportion of women authoring in leading medical journals varied by the country where the research had been conducted (Fig. 1G). Female authors were least common in publications originating from South Korea, Japan, Singapore, Germany, and Austria, whereas they were most frequent from South Africa and India. The gender gap was more pronounced for second last and last authors with 24.8% female authors in median across countries compared to first and second authors with 34.5% female authors (p<0.001). Gender underrepresentation was highest in South Korea with 0.0% (0/28), Japan with 0.9% (1/106) and Austria with 9.1% (3/33) female last authors.

Next, we examined longitudinal national trends in gender representation. The gender-associated difference in publication number decreased rapidly in UK, Canada, and Belgium both for first and last authors (Supplementary Fig. 1A-B). Yet, the gender gap remained evident at the end of the follow-up in many countries, such as in the USA, UK, and China, and particularly at the last author level. In opposite to the general trend, the gender gap rose for last authors of publications from Israel and both for first and last authors from China. Collectively, these findings imply that the longitudinal trend could guide in customizing national measures to mitigate gender underrepresentation.

### Collaboration scope varies by gender and is associated with publication and citation count

Next, we interrogated the significance of collaboration scope over gender-related differences in citation count. The number of co-authors (*R* 0.46, p<0.001) correlated with citation count. Male first and last authors published in average with one author more than their female colleagues (Table 1). To measure the independent impact of author count and gender on citations, we applied a linear regression model using these and their interaction term as covariates. In parallel, we examined the association of gender and citations in subgroups of categorized authorship count. Both approaches revealed that author count and gender are independent but weak predictors of future citation count (Supplementary Table 1, Supplementary Fig. 2).

When examining the temporal evolution of authorship patterns, we observed a stable median increase of 9 additional authors per article during the 10-year follow-up (Supplementary Fig. 3A). Publications with more than 11 authors doubled in that time and these accumulated faster citations per article (2.99 citations/article/year vs. 1.19 citations/article/year; Fig. 2A-B). The inclination was also 55.3% steeper for men as first author and 94.1% for men as last authors during the 10-year follow-up compared to corresponding female authors (Supplementary Fig. 3B). Instead, the slope between first and last authors did not differ when comparing separately women and men (Supplementary Fig. 3B).

**Figure 2.**
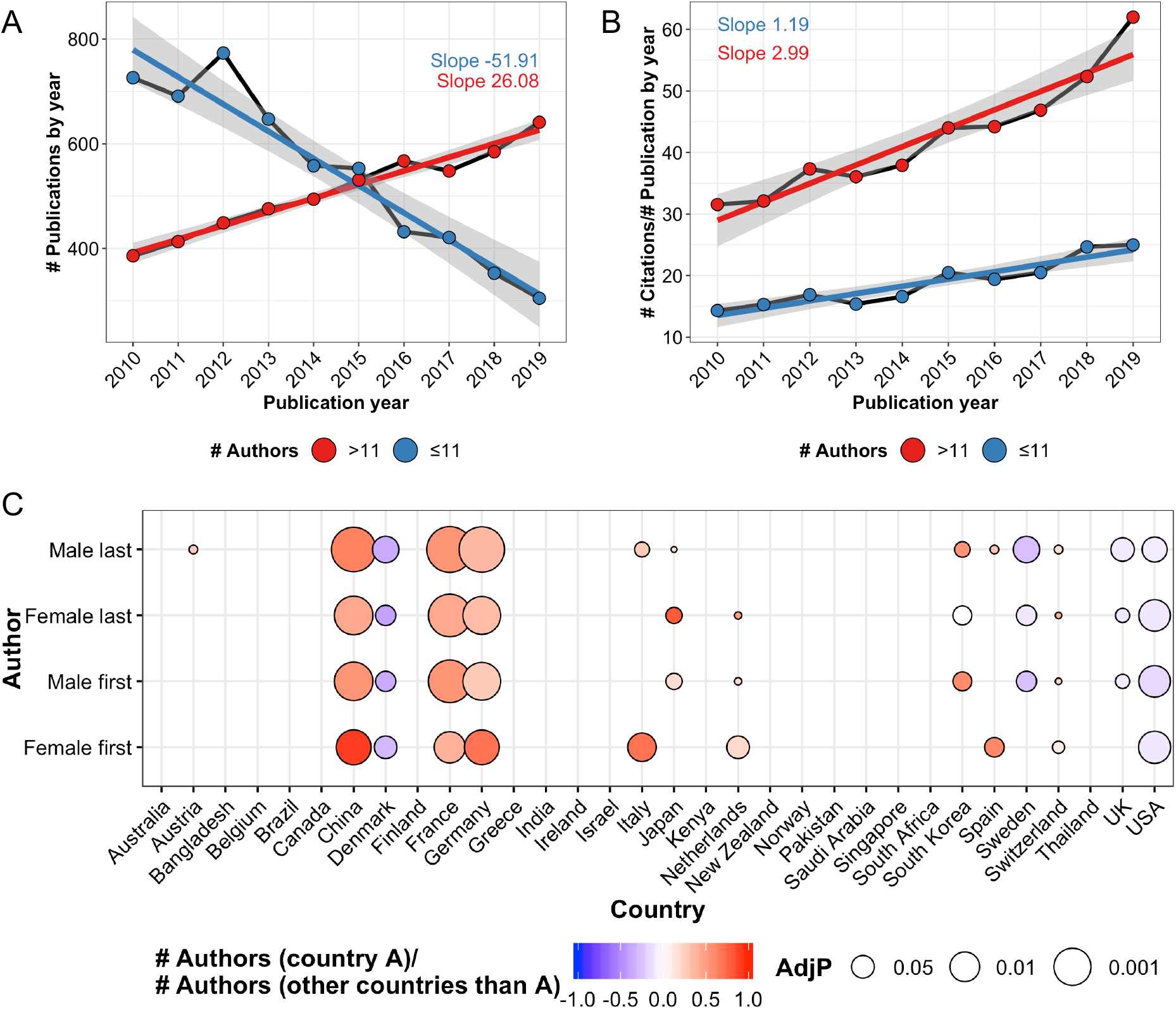
Scope of collaboration by genders. (A) Line plot and fitted linear regression for the number of publications and (B) yearly-averaged citations per publication by their publishing year and number of authors. (C) Balloon plot on the association of the first and last author gender distribution by the affiliation nationality of the corresponding author. The balloon color reflects the number of authors by country (column) compared to all other countries and the balloon size the adjusted p value of that comparison.

To study national differences in collaboration scope, we compared the number of authors for publications by gender and by country (Fig. 2C). While the number of authors did not differ for most countries, almost all variation in the number of authors were related to the geographical origin of the research. Publications from USA, UK, Denmark, and Sweden shared the least authors (Fig. 2C). On the opposite, publications from France, Germany, China, and South Korea were associated with more numerous authors/article (Fig. 2C). Together, the findings emphasize that the geographical origin of the research has a more pronounced association with collaboration scope than gender.

### The thematic and journalistic disparity between female and male medical researchers

Previous reports have suggested differences in funding, mentorship training, and household and caregiving responsibilities between women and men to account for the gender gap in medical publishing^2,11^. We hypothesized whether the areas of research would differ by gender. We observed a clear correlation between keyword-associated citation frequency and the proportion of male first (*R* 0.40, p<0.001) and last authors (*R* 0.40, p<0.001; Fig. 3A). Keywords associated with highest publication citations were related to phase II-III trials, oncology, immunotherapy, chemotherapy and antibody-based therapy, and were enriched with publications authored by men, especially at the senior author level. On the contrary, the 20 keywords associated with least citations were predominantly associated with female authorship, notably in the context of first authorship (Fig. 3A). The keywords covered healthcare-related themes such as patient involvement, insurance, quality-of-care, and access.

**Figure 3.**
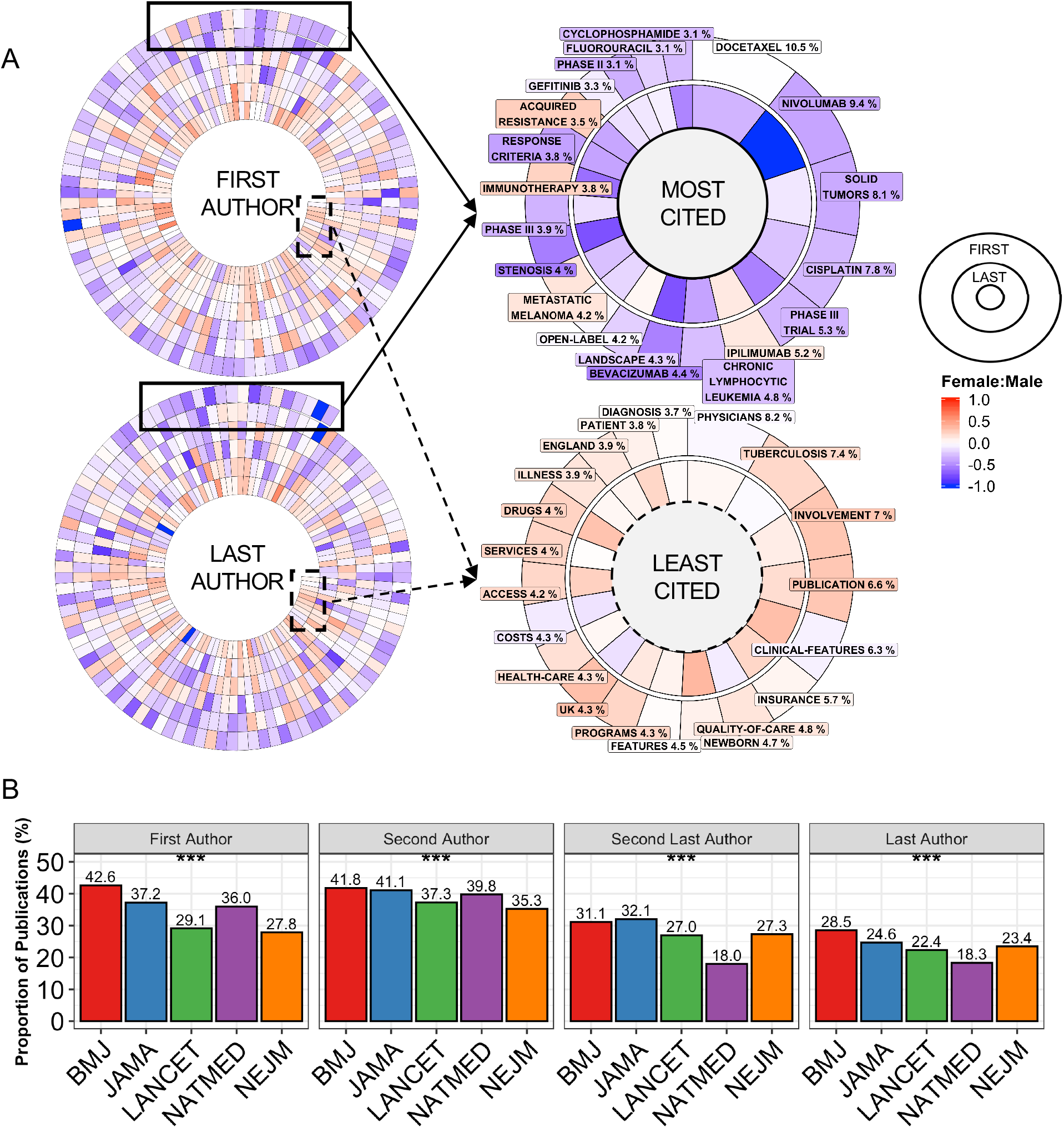
Gender gap by subfields and journals. (A) Left side spiral plots illustrate the keywords occurring ≥20 times and arranged by their citation impact starting from the outside layer (most cited) towards the inner core (least cited). Right side plots show the proportion (size of the bar) of the most and least cited keywords. The color of the bars in all spiral plots illustrate the gender balance. (B) Bar plot illustrating the proportion of publications in medical journals originating from different countries.

Beside gender-associated distinctions in research subfields, we sought to understand whether publications sharing the same keyword would differ by their accumulated citation count. Out of 583 keywords, we included 579 for comparing first author and 582 for last author gender comparison as these keywords were used by both genders. For first authors, 78/579 compared to 5/579 keywords resulted in higher citation count (adjusted p<0.05) when employed by men vs. women, respectively. For last authors, the corresponding proportion was 32/582 for men and 8/582 for women (adjusted p<0.05). Collectively, the findings indicate that within the same fields of research, publications authored by women accumulate fewer citations compared to publications authored by men.

To conclude, we investigated the proportion of female-authored publications by journals. The absolute difference in the proportion of first authors between the five journals was 14.8% for first and 10.2% for last authors (Fig. 3B). Female first and second authors were least frequent in articles published in *NEJM* and *Lancet*, whereas second last and last authors were least common in *NatMed*. According to this analysis, *BMJ* and *JAMA* were the most representative journals considering all four authorship positions.

## DISCUSSION

Available bibliometric data can reveal novel information on the equity and diversity of scientific research. Longitudinal data permits studying temporal trends, which can be crucial for monitoring purposes. Here, we presented publication disparity in five leading medical journals between 2010-2019. The study analyses can be easily replicated for any journals and any time period using data available at the Web of Science’s database and codes published with this study.

Gender underrepresentation in scientific publishing has been documented earlier. Women tend to obtain university-level degrees more commonly than male in OECD countries (47% vs. 32%)^12^. However, female doctoral students submit and publish less than their male colleagues^2,3^. The difference is largest in natural/biological sciences and engineering and has been hypothesized to result of unevenly distributed resources and support^3^. Gender disparities in publications have been shown to correlate with future academic rank signifying sustained impact on professional careers^13^, salary and job satisfaction^14^.

Our findings in leading medical journals are in line with previous observations. For the first time to our knowledge, we report that the inequity is country-specific. While the distinction is visible across first, second, second last and last author positions, the gender gap is most pronounced at the senior author level. In particular, publications with a corresponding author in Germany, Austria, Japan, South Korea, and Singapore had the fewest female authors. The finding implicate that the bias likely arises from national and cultural factors rather than editorial or peer-review processes, which had previously indicated mixed results^15,16^. Moreover, the gender gap in first and last authorships has steadily declined during the last decade in most countries, but the progress has been less evident or even contrary in some countries such as China and Israel. The gender disparity has decreased more rapidly in last authorships possibly reflecting a response to the more pronounced gender underrepresentation compared to first authors or dynamics in generation transition.

Gender underrepresentation was observable in all top medical journals. However, female first and second authors were least frequent in articles published in *NEJM* or *Lancet*, whereas second last and last authors were least common in *NatMed*. The findings are in line with a previous study examining gender representation as first authors using 4 out of 5 similar journals during 1994-2014^17^. The trend was replicated also in an article examining author disparity in leading medical journals during the COVID-19 pandemic^11^. In that study, no difference was found when examining first and last author gender of COVID vs. non-COVID-related research^11^. Confirming our longitudinal findings, the proportion of female first (36.2% vs. 33.6%) and especially last (29.5% vs. 23.4%) authors has increased in 2020 compared to our data covering 2010-2019. Similar findings were not evident between our and the earlier study covering 1994-2014 emphasizing that the gender representation has started to improve only recently.

By comparing research keywords, first and especially last male authors tended to publish clinical trials and oncology-associated studies, which accumulated highest citation counts. The difference in research focus has not been demonstrated before and likely accounts for some of the journal-specific gender disparities. However, our data also indicated that publications first or last-authored by men tend to accumulate more citations implying that differences in research fields explain only limited variability.

In line with results of our study, articles with a woman researcher as first or last author have been shown to accumulate fewer citations^18^. While medical research was not included, a previous study using 1.5 million interdisciplinary papers in 1779-2011 has indicated that male first authors tend to self-cite 56% themselves more commonly than their female colleagues, and even more during the last decades^19^. According to a recent preprint, males were more commonly quoted in Nature science journalism, which could skew the recognition and future citation probability of publications by gender^20^.

Between 2010-2019 articles the number of authors increased by nine reflecting a fundamental change in research towards larger collaboration and building consortia. This correlated with higher citation frequency per article. While the general citation rate increased in the top medical journals, the rate was over 2.5 times faster in articles with more collaborators. We found that publications with male researchers had more authors in concordance with a previous study^21^. In our study, both author gender and number of collaborators were independent but weak predictors of future citation rate.

Computational gender prediction could be a source of error as some names can be used by both women and men, especially in different countries. The name-based analysis may also misclassify authors with non-binary genders. However, previously reported gender-related bibliometric observations were in line with our findings indicating that the gender prediction based on authors’ first names provided realistic results. Inclusion of gender ethnicity and career stage were unavailable but could be important factors to further study gender representation.

In summary, this computational audit indicated that gender disparity in medical research is country-specific, partly related to distinct research focus and more evident at the senior researcher level. The findings might reflect available financial resources and research support. The analysis also highlighted that the gender gap is decreasing with country-dependent variability.

## Data Availability

Codes and a 100-row data example are available at https://github.com/obruck/International-Research-Impact. Raw data can be downloaded from Clarivate Web of Science, with instructions provided in the Github repository.

## ADDITIONAL INFORMATION

## ACKNOWLEDGMENTS

The author wishes to thank Susanna Lallukka-Brück for her patience, understanding, and insightful comments. The author is grateful for Olli Dufva and to the members of the Hematoscope Lab for discussion and comments. Certain data included herein are derived from Clarivate Web of Science. © Copyright Clarivate 2022. All rights reserved.

## AUTHORS’ CONTRIBUTIONS

Conception and design; Collection and assembly of data; Data analysis; Manuscript writing; Manuscript editing; Data interpretation; Final approval of manuscript: O.B.

## ETHICS APPROVAL AND CONSENT TO PARTICIPATE

The study complied with the Declaration of Helsinki.

