## Supplementary Data for "The Gender Gap in Leading Medical Journals - a Computational Audit"

Supplementary table 1. Linear regression models with log10-transformed year-averaged citation count.

| Model | Model covariates | Covariate | Coefficient estimate | $R^2$ | P-value |
|---|---|---|---|---|---|
| 1 | Cov1 | Cov1 | 0.0012, p<0.001 | 0.026 | p<0.001 |
| 2 | Cov2 | Cov2 | 0.099, p<0.001 | 0.0061 | p<0.001 |
| 3 | Cov3 | Cov3 | 0.083, p<0.001 | 0.035 | p<0.001 |
| 4 | Cov1+ Cov2+Cov1*Cov2 | Cov1 | 0.0030, p<0.001 | 0.036 | p<0.001 |
| 4 | Cov1+ Cov2+Cov1*Cov2 | Cov2 | 0.12, p<0.001 | 0.036 | p<0.001 |
| 4 | Cov1+ Cov2+Cov1*Cov2 | Cov1*Cov2 | -0.0018, p<0.001 | 0.036 | p<0.001 |
| 5 | Cov1+ Cov3+Cov1*Cov3 | Cov1 | 0.0011, p<0.001 | 0.034 | p<0.001 |
| 5 | Cov1+ Cov3+Cov1*Cov3 | Cov3 | 0.068, p<0.001 | 0.034 | p<0.001 |
| 5 | Cov1+ Cov3+Cov1*Cov3 | Cov1*Cov3 | 0.00073, p<0.001 | 0.034 | p<0.001 |

Abbreviations: Cov1, Number of authors; Cov2 = First author gender (male); Cov3 = Last author gender (male); Cov1*Cov2, interaction of Cov1 and Cov2; $R^2$, adjusted R-squared

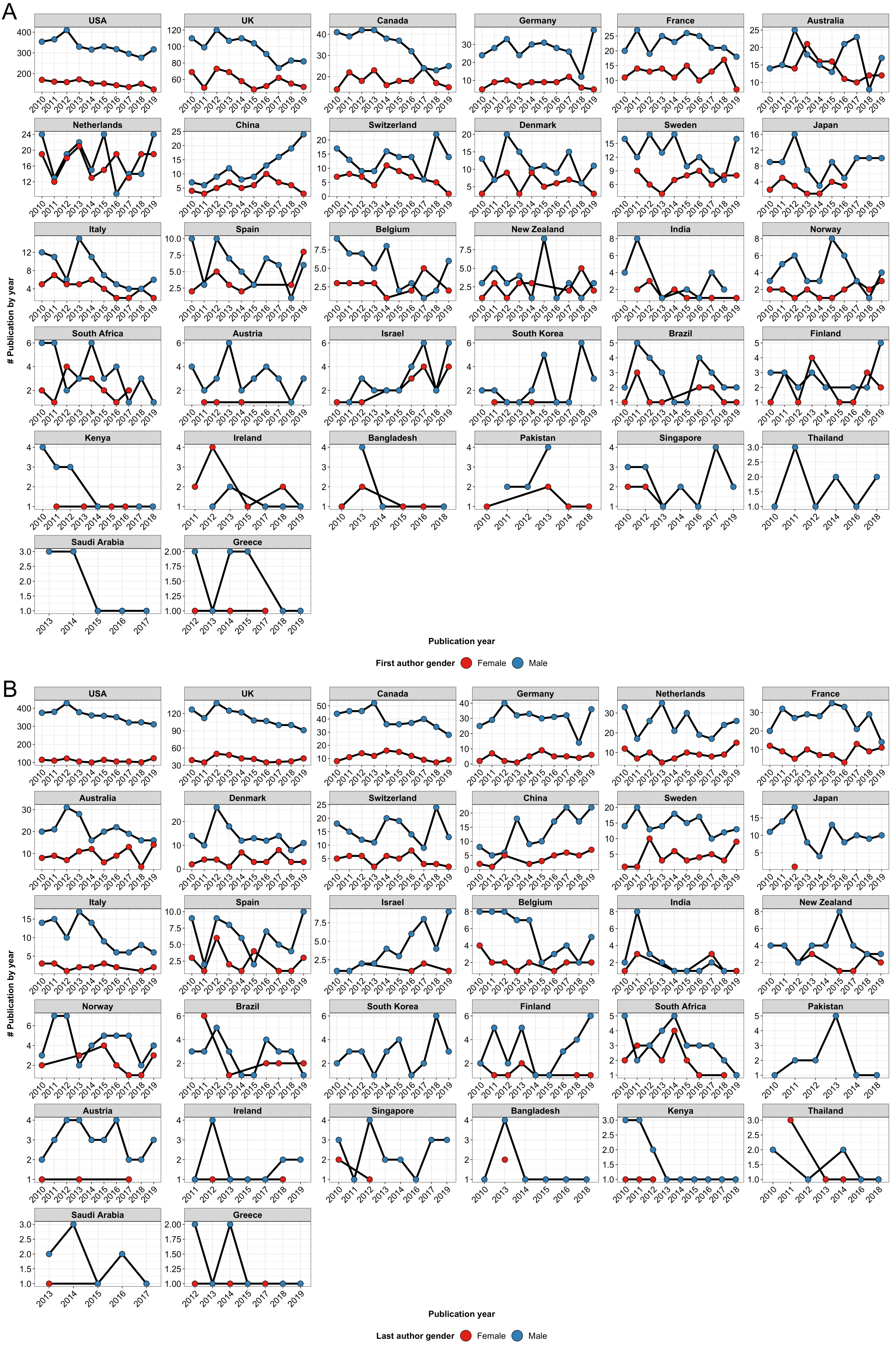


Supplementary Figure 1. Line plot illustrating the number of publications by their publishing year from the top 32 most productive cities based on the gender of the (A) first and (B) last author.

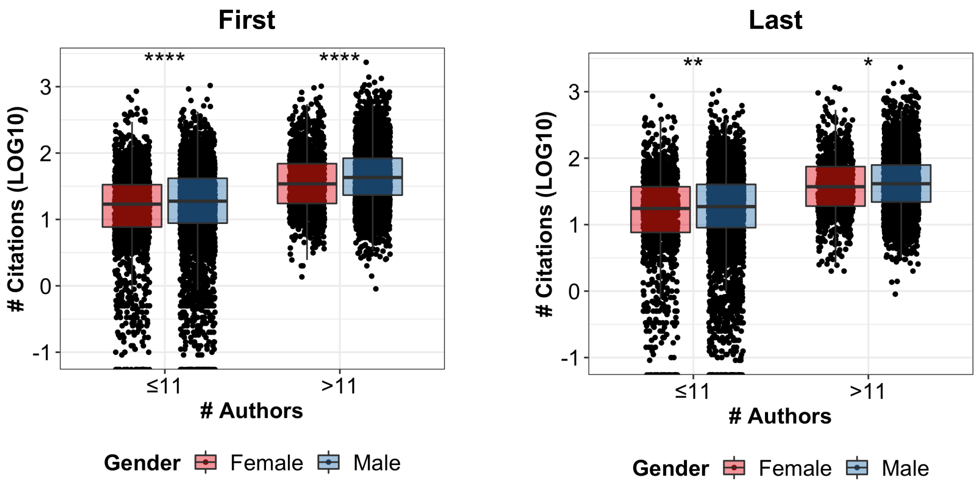


Supplementary Figure 2. Box plot illustrating the yearly-averaged citations (LOG10-transformed) per publication. Citations are compared by the number of authors (median 11 authors) and gender.

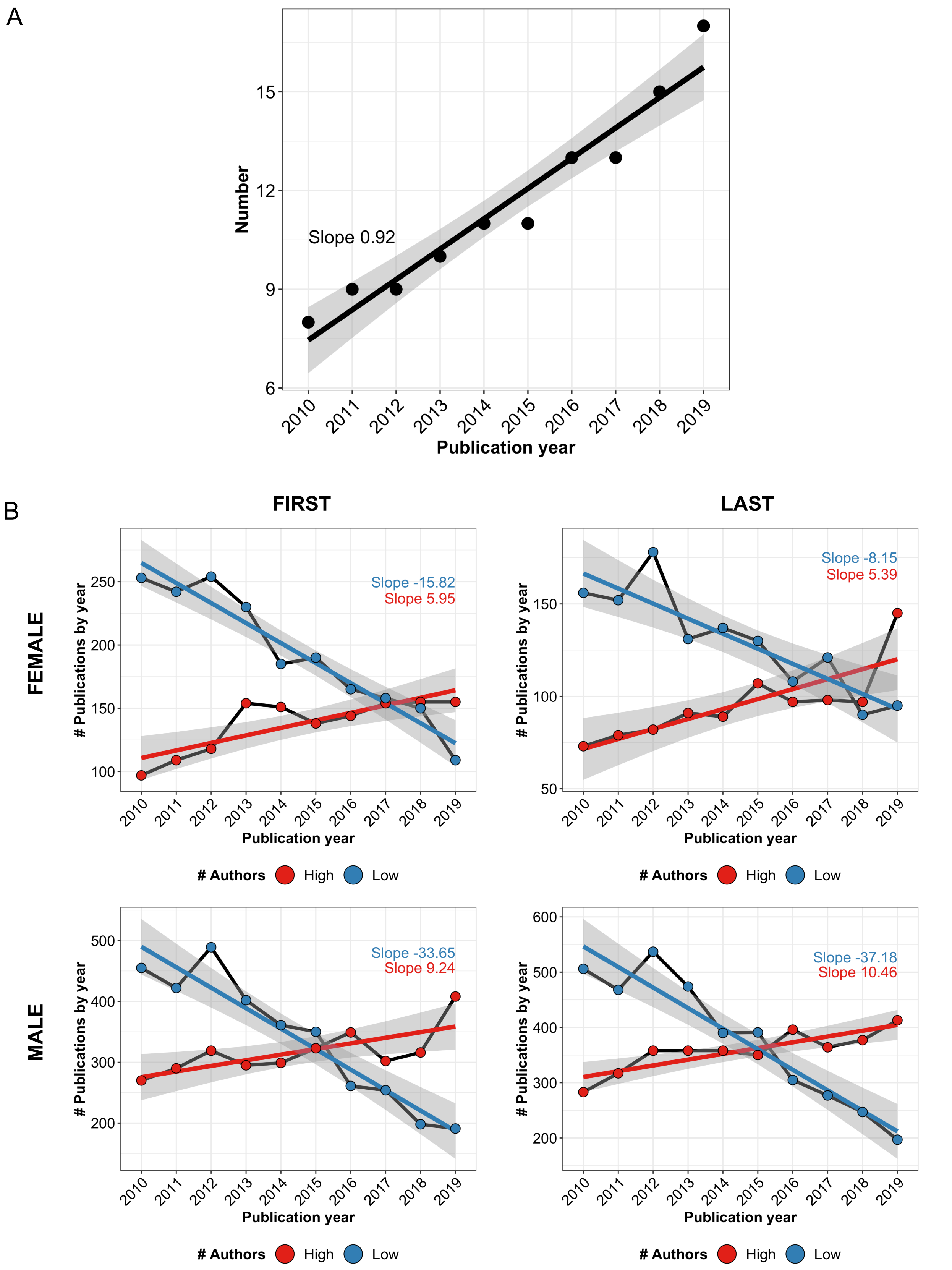


Supplementary Figure 3. (A) Fitted linear regression for the number of authors in publications by their publishing year. (B) Line plot and fitted linear regression for the number of publications by their publishing year and number of authors. The panel is divided by first and last author gender.
